# Epidemiology, clinical features, and visual outcomes after intraocular foreign body removal: an IRIS® Registry (Intelligent Research in Sight) Analysis

**DOI:** 10.1101/2025.09.08.25335244

**Authors:** Ariel Yuhan Ong, Eric A Goldberg, William C Kearney, Connor Ross, Caroline Awh, David A Merle, Siegfried K Wagner, Robbert R Struyvven, Pearse A Keane, Tobias Elze, Joan W Miller, Alice Lorch, Lucia Sobrin, Ines Lains, the IRIS® Registry Analytic Center Consortium

**Affiliations:** Institute of Ophthalmology, University College London, United Kingdom; Moorfields Eye Hospital NHS Foundation Trust, London, United Kingdom; NIHR Moorfields Biomedical Research Centre, London, United Kingdom; Department of Ophthalmology, Massachusetts Eye and Ear, Harvard Medical School, Boston, Massachusetts USA

**Keywords:** Intraocular foreign body, Outcomes, IRIS Registry, Pars plana vitrectomy, Retinal detachment

## Abstract

**Purpose:** To describe the epidemiology, clinical features, and visual outcomes of intraocular foreign body (IOFB)-associated eye injuries.

**Design:** Retrospective multicenter cohort study utilizing data from the American Academy of Ophthalmology IRIS® Registry (Intelligent Research in Sight).

**Subjects:** Eyes that underwent IOFB removal between January 2016 and October 2024.

**Methods:** Sociodemographic information, clinical features at presentation, primary surgical procedures and postoperative complications were summarized. Multivariable linear mixed-effects regression models were employed to investigate predictors of visual outcomes up to 18 months post-IOFB removal.

**Main outcome measure:** Epidemiology (including annual incidence rates and associated factors) and clinical characteristics; predictors of visual acuity (VA) up to 18 months post-IOFB removal.

**Results:** A total of 4784 eyes (4684 patients, 70.3% male) with a median age of 55 years at presentation (interquartile range 36-70) were identified over the study period. Mean annual incidence was estimated at 2.32 per 100,000 patient-years (95% CI 2.12-2.52) and was independently associated with male sex, race, and rural residence. The most common complications at presentation were retinal detachment (12.5%), cataract (10.5%), vitreous hemorrhage (7.9%), and endophthalmitis (3.9%). Median VA at presentation was 1.24 logMAR (IQR 0.30-2.30). A significant improvement in VA was seen only from month two post-IOFB removal (−0.38 logMAR, 95%CI −0.41 to −0.34), with further minor improvements up to month 18 (−0.59 logMAR, 95%CI −0.69 to −0.48). After adjusting for relevant covariates, Black or African American race and presence of endophthalmitis, retinal detachment or hyphema at baseline were associated with worse visual outcomes. Subgroup analysis of patients with pre-IOFB VA found that improvement was attenuated for people with pre-IOFB VA worse than 1.0 logMAR.

**Conclusions:** These findings offer a real-world benchmark for post-IOFB visual trajectories and outcomes, and may support clinicians in prognostication and patient counselling. Further research is needed to investigate the underlying drivers of observed racial disparities to inform equitable care.

---

Open globe injuries (OGI) involving intraocular foreign bodies (IOFB) are ophthalmic emergencies with the potential for severe and lasting visual impairment. Despite their clinical significance, there remains a lack of contemporary, population-level data on the sociodemographic profiles, clinical presentation, and long-term visual outcomes associated with these injuries.

Existing literature on the topic is limited in scope. Most published studies to date predominantly focus on OGI, and reports on IOFBs mostly comprised small sample sizes and single-center settings, with a dearth of longitudinal follow-up.^1–9^ The available literature includes data largely collected from previous decades, which may not fully capture the impact of modern surgical techniques and treatment protocols (including antimicrobial prophylaxis practices), thus limiting their utility for guiding prognosis and clinical management. In addition, while these studies were conducted across a diverse range of countries, robust data from high-income countries are lacking.

While sociodemographic factors such as race and ethnicity, income, education, or geographic disparities are increasingly recognized to be associated with health outcomes across surgical and trauma care,^10^ how these factors intersect with treatment outcomes has not been as well characterized. Better understanding of these dimensions may be helpful in guiding clinical decision-making and patient counselling, while informing the design of targeted public health initiatives aimed at prevention and management of IOFB-related injuries and improving health equity.

This study aimed to address these research gaps using a large multicenter clinical registry to provide a better understanding of contemporary presentation, management and factors influencing visual prognosis in routine clinical practice. In particular, we assessed the 1) epidemiological and sociodemographic characteristics of patients presenting with IOFB injuries requiring surgery in the American Academy of Ophthalmology IRIS® Registry (Intelligent Research in Sight), a large United States (US)-based dataset of over 80 million patients, 2) clinical presentation and procedures performed around the time of presentation, and 3) the long-term visual outcomes following IOFB removal and their predictors.

## Methods

### Study design and data source

This was a multicenter retrospective observational cohort study of eyes with IOFBs treated in the US. The study period spanned January 2016 to October 2024. The study utilized data from the IRIS Registry, one of the largest specialty-specific clinical data registries in the world. The IRIS Registry captures structured electronic health record (EHR) data from a broad network of ophthalmology practices encompassing diverse geographic regions, subspecialties, and practice settings across the US. Details of the EHR data extraction process, data fields, and distribution of practices contributing data has been published previously.^11^ This version of the database was frozen on October 31, 2024.

The IRIS Registry is a centralized data repository and reporting tool that can be used for research purposes. This does not constitute human subject research because data in the IRIS Registry is de-identified, and the investigator does not have access to study identifiers. Therefore, institutional board review and informed consent are not required. This study adheres to the Declaration of Helsinki.

### Study population

Clinical diagnoses were identified using International Classification of Diseases (ICD) 10^th^ Edition and Systematized Nomenclature of Medicine Clinical Terms (SNOMED-CT) codes, while procedures were identified using Current Procedural Terminology (CPT) codes, allowing for consistent classification across institutions and time.

#### Inclusion criteria

Eyes with CPT codes for IOFB removal (65235, 65260, 65265) were identified, and the first-ever instance of this code was assigned as the baseline date.

#### Exclusion criteria

Eyes with CPT codes for retained lens fragments following cataract surgery, glaucoma procedures (e.g. goniotomy, insertion of aqueous shunt), removal of implanted foreign materials (e.g. removal of aqueous shunt), or revision thereof (e.g. intraocular lens (IOL) repositioning, aqueous shunt revision) within 3 days of IOFB removal were excluded. This was a conservative approach designed to minimize potential errors from miscoding. The full list of codes used is provided in Supplementary S1 (available at https://www.aaojournal.org).

### Variables of interest and data handling

Sociodemographic information including age, sex, race and ethnicity, median income, high school graduation percentage, operating practice geographic location, and urban/rural status were extracted. Operating practice geographic location was converted from state to region based on US Census classifications. Median income and high school graduation percentage are based off the 2021 American Community Survey results for the patient’s recorded zip code.

Clinical characteristics of interest included IOFB extraction method (magnetic, non-magnetic, or not recorded (NR)), whether they had an IOFB in one or both eyes, IOFB location, and contemporaneous ophthalmic procedures recorded at baseline or up to 3 days prior to this, to account for the possibility of delayed primary repair and/or sequential IOFB removal. These data fields were extracted and summarized from a list of CPT codes performed at baseline. Complications at presentation were recorded up to 3 days prior to IOFB removal for the same reasons and were detailed separately for ‘early complications’ (1-6 days) and ‘later complications’ (7 days onwards). The list of complications was informed by a comprehensive literature search, and were extracted using pre-specified ICD-10 codes listed in Supplementary Table 2 (available at https://www.aaojournal.org).

Logarithm of the minimum angle of resolution (logMAR) visual acuity (VA) was used for analysis. Any VA recorded in Snellen fractions was converted to the appropriate logMAR equivalent using a standard conversion table for the IRIS Registry (-log(Snellen fraction), log base 10). All VA measures reported utilize best documented distance visual acuity (BDVA) for a given date and eye. Presenting VA was similarly defined as being the mean of BDVA recorded at baseline or up to 3 days prior to this. VA analyses were restricted to the first 18 months post-IOFB removal for maximal robustness. For the small subset of patients with pre-IOFB VA measurements between 1-12 months prior to baseline, eyes were categorized into one of 3 categories based on their mean pre-IOFB BDVA : −0.3 to 0.5 logMAR, 0.5 to 1.0 logMAR, and >1.0 logMAR.

### Statistical analysis

Categorical data were described with number and proportion, and continuous data with the median and interquartile range (IQR) after confirming a non-normal distribution with the Shapiro-Wilk test and Q-Q plots.

Annual incidence rates (per 100,000 patients) were calculated as the number of patients undergoing IOFB removal in that year divided by the number of patients recorded in the IRIS Registry over that same period with 95% confidence intervals (CI) estimated via the Poisson distribution. Incidence calculations were performed at the patient level, with bilateral same day cases counted as a single case. Comparison of incidence rates over time was conducted using the exact Poisson test. Multivariable logistic regression models were employed to estimate the odds ratios (OR) and 95% CI for factors associated with IOFB removal; this comprised sociodemographic covariates such as age, sex, race and ethnicity, median income, and urban/rural status.

Two linear mixed-effects models were then employed to assess predictors of post-IOFB VA, incorporating random intercepts at the eye level to account for the non-independence of repeated measurements. For the small number of bilateral cases, one eye was selected at random for VA modelling. A random slope for time was included to model potential heterogeneity in individual VA trajectories over time. Sociodemographic factors and clinical features present at baseline were included as covariates. The first model examined the change in post-procedure VA at each month post-IOFB compared to baseline to map the trajectory of VA change. The second model was a subgroup analysis of patients with pre-IOFB VA measurements to evaluate the possibility of poorer visual outcomes secondary to pre-existing ocular comorbidities in this cohort.

To test the robustness of the findings under the potential influence of outliers, a sensitivity analysis was performed using a mixed-effects quantile regression model focused on the median rather than the mean. This is because the median is less sensitive to extreme values (e.g. very poor or very good VA) and does not assume normality of the residuals. All *P*-values for model coefficients were adjusted using the Benjamini–Hochberg procedure to control the false discovery rate (FDR) across multiple comparisons.

Data analysis was performed using R version 4.4.2 (R Core Team, 2024). Predictors were considered statistically significant at a level of *P*<0.05.

## Results

### Epidemiology

A total of 4784 eyes (4684 patients) met the criteria for inclusion within the study period. The mean annual incidence was 2.28 (95% CI 2.09-2.49) per 100,000 patient-years. There was a small, gradual decrease in annual incidence from 2017 to 2023 (from 2.84 per 100,000 patient-years in 2017 to 1.95 in 2023, *P <* 0.001). Although this trend continues into 2024, data for the year are incomplete (up to October 2024) (Table 1).

Table 2 summarizes the sociodemographic characteristics of the overall study cohort. The median age was 55 years (IQR 36-70), with a total of 172 pediatric patients aged under 18 (3.7%, 176 eyes). Most patients were male (70.3%), White (59.6%), and resided in urban areas (82.6%). They were treated in practices that were predominantly based in the South (42.6%). The majority resided in areas with a median income of USD 35,000-74,999 (52.0%), broadly aligning with the national average.

After adjustment for relevant covariates, factors independently associated with an increased odds of IOFB removal included male sex (OR 3.44, 95% CI 3.22-3.67), Other race (OR 1.61, 95% CI 1.16-2.23) or Hispanic/Latino ethnicity (OR 1.49, 95% CI 1.35-1.64), and residence in a rural area (OR 1.65, 95% CI 1.52-1.79). Older age (OR 0.88 per decade over 65, 95% CI 0.87-0.90) and a median income category of USD 75,000-149,999 (OR 0.80, 95%CI 0.74-0.86) were protective.

Demographic trends across time are reported at the eye level due to a small proportion (33, 0.66%) of patients sustaining IOFBs in their fellow eye sequentially. While there were minor year-to-year fluctuations in absolute percentages, the majority of demographic categories (such as male sex, White ethnicity, urban residence, Southern geographic region, and mid-range income bracket) remained consistent with the overall cohort profile throughout the study period (Supplementary Table 3, available at https://www.aaojournal.org).

### Clinical characteristics and procedures at baseline

Clinical characteristics and procedures performed at baseline are summarized in Table 3. Most cases were unilateral (96.9%). The majority of IOFBs were located in the posterior segment (51.8%), followed by the anterior segment (46.6%), with a small percentage affecting both (1.6%). IOFB extraction methods were not recorded in cases occurring in the anterior segment (46.6%); where this was recorded for IOFBs in the posterior segment, non-magnetic extraction was most common (40.9%), followed by magnetic (12.1%), and both techniques (0.3%).

A B-scan ultrasound was performed in 9.0% of patients at baseline. Procedures conducted contemporaneously with the IOFB removal included pars plana vitrectomy (PPV) with or without retinal detachment (RD) repair (42.5%), repair of anterior segment laceration(s) (17.8%), anterior chamber (AC) washout (14.6%), and lens extraction (11.5%). Intravitreal injections (drug unspecified) were performed in 3.7%, while intravitreal antibiotic injections (ceftazidime/vancomycin) were recorded in an additional 1.8%. Of note, 1367 eyes (28.6%) had ≥2 additional procedures (Table 3).

The most common complications at baseline were RD (12.5%), cataract (10.5%), vitreous hemorrhage (7.9%), and endophthalmitis (3.9%) (Table 4). An additional 14.3% of eyes developed RD over the course of their follow-up at a mean of 9.4 months (median 2.4, IQR 1-9 months) at an incidence of 8.4 per 100 eye-years (95% CI 7.8-9.0) over a median follow-up of 16.9 months (IQR 4.2-43.9).

### Visual outcomes post-IOFB removal

VA at presentation was available for 1941 eyes. Median VA at presentation was 1.24 logMAR (IQR 0.30-2.30) (approximately 20/400 Snellen), with 54.5% (1058/1941) having a vision of 1.00 logMAR (20/200 Snellen) or worse. Of the patients in this cohort, 12.2% (237/1941) presented with light perception (LP) and 1.3% (26/1941) with no light perception (NLP).

### Predictors of changes in VA from baseline

VA data was available for longitudinal analysis in 1376 unique eyes, requiring one measure at baseline and at least two more within 18 months post-op. Changes in VA from baseline post-IOFB removal featured a distinct pattern of VA recovery, with no significant change at month 1 (β 0.01, 95% CI −0.02-0.04), a statistically significant improvement of 0.38 logMAR on average by month 2 (95% CI −0.41 to −0.34), and a gradual improvement to 0.59 logMAR that was maintained to month 18 (95% CI −0.69 to −0.48) (Figure 1).

**Figure 1:**
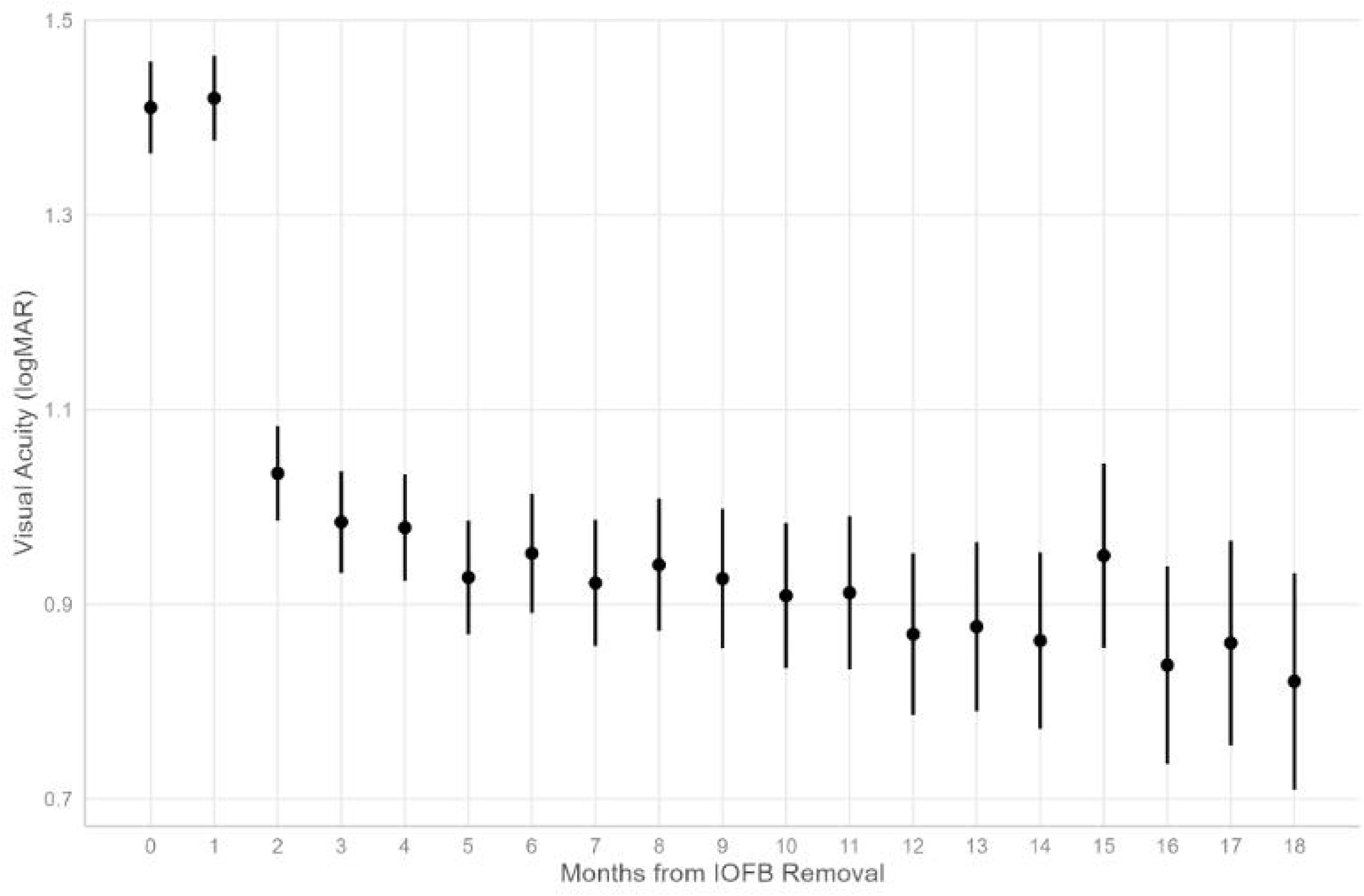
Longitudinal trajectory of visual acuity up to 18 months after sustaining an intraocular foreign body injury (IOFB). The error bars indicate the 95% confidence interval.

**Figure 2:**
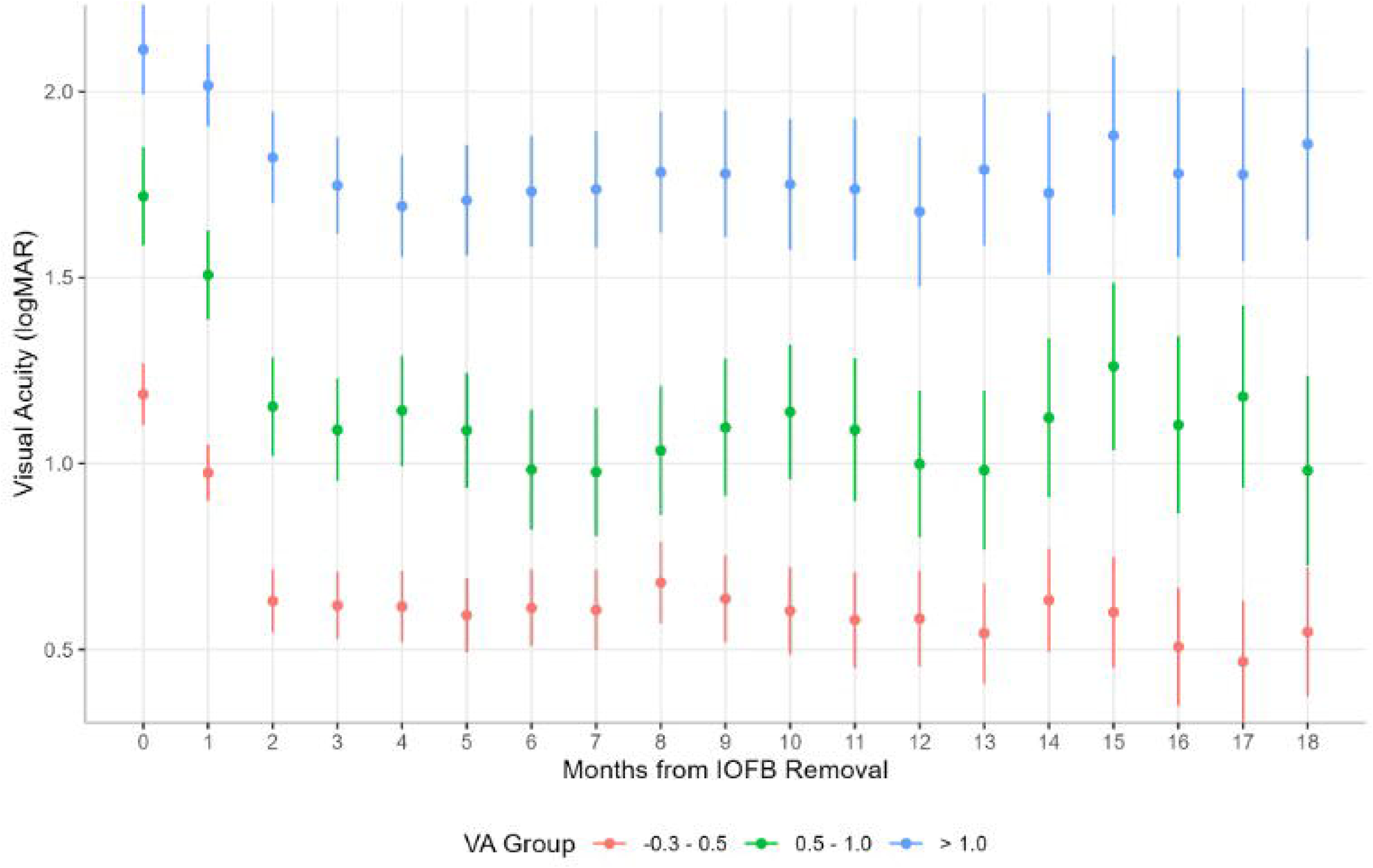
Longitudinal trajectory of visual acuity up to 18 months after sustaining an intraocular foreign body (IOFB) injury, stratified by pre-IOFB visual acuity (logMAR −0.3 to 0.5, 0.5-1.0, and >1.0). The error bars indicate the 95% confidence interval.

After adjusting for baseline clinical characteristics, several factors were significantly associated with poorer VA outcomes (Supplementary Table 4, available at https://www.aaojournal.org). Baseline complications including endophthalmitis (β 0.70, 95% CI 0.56–0.84), hyphema (β 0.74, 95% CI 0.55–0.93), RD (β 0.35, 95% CI 0.22– 0.48), and vitreous hemorrhage (β 0.23, 95% CI 0.10–0.36) were strong predictors of worse overall vision. Compared to anterior segment-only injuries, IOFBs affecting the posterior segment (β 0.25, 95% CI 0.15–0.35) or both anterior and posterior segments (β 0.58, 95% CI 0.07–1.09) were also associated with significantly worse VA.

In terms of sociodemographic characteristics, self-identifying as Black or African American was independently associated with a worse visual outcome (β 0.35, 95% CI 0.17–0.52) even after controlling for all clinical and other socioeconomic variables. No statistically significant association was found for other socioeconomic factors, including median household income or urban-rural status. The interaction between race and median income was not significant as well. The final model explained a substantial portion of the variance in VA outcomes (marginal R² = 0.200, conditional R² = 0.750).

### Sensitivity analysis

Sensitivity analysis was conducted using a mixed-effects quantile regression model to assess predictors of the median visual outcome and provide a model robust to outliers. Median logMAR VA improved over time, with a stable improvement of 0.26-0.36 logMAR units from month 2 onwards compared to baseline (all adjusted *P*<0.001). Compared to the earlier analysis, only three clinical factors remained associated with a poorer median VA – the presence of endophthalmitis (β 0.42, 95%CI 0.05-0.79]) and RD (β 0.418, 95%CI 0.17-0.67), and IOFB affecting both anterior and posterior segments (β 0.50, 95% CI 0.07–0.93) at baseline. In this median-focused model, sociodemographic factors and presence of other complications at presentation were not significant predictors of VA (Supplementary Table 5, available at https://www.aaojournal.org).

### Subgroup analysis of eyes with pre-IOFB VA

A subgroup analysis comprising 625 eyes with at least one pre-IOFB VA measure (between 1 to 12 months pre-IOFB) and post-IOFB VA reading was performed. In general, this subgroup was older, had a greater proportion of White patients, more female representation, and a higher proportion of baseline complications compared to the overall cohort (see Supplementary Table 6 for cohort characteristics, available at https://www.aaojournal.org). For this analysis, eyes were stratified into three groups based on their pre-IOFB VA: good (−0.3-0.49 logMAR), moderate (0.5-1.0 logMAR), and poor (>1.0 logMAR). Pre-IOFB VA was the strongest predictor of post-IOFB visual outcomes. While VA generally improved over time for all three groups, a significant interaction effect demonstrated that this improvement was attenuated for the group with poor pre-IOFB VA (>1.0 logMAR).

Consistent with previous findings, in this subgroup, several other clinical factors were associated with worse visual outcomes, including the presence of endophthalmitis (0.73, 95%CI 0.56-0.91), RD (0.52, 95%CI 0.31-0.73), vitreous hemorrhage (β 0.39, 95%CI 0.14-0.64), and hyphema (β 0.37, 95%CI 0.16-0.58). Patients identifying as Black or African American (β 0.31, 95%CI 0.12-0.50) and Other (β 0.25, 95%CI 0.08-0.41) also had worse visual outcomes. IOFB location was not found to be a significant predictor of visual outcomes in this model (Supplementary Table 7, available at https://www.aaojournal.org).

## Discussion

In this large longitudinal IRIS Registry study of eyes affected by IOFB injuries, we have mapped the epidemiology of IOFB from 2016-2024, identified a distinct trajectory of visual recovery and delineated key clinical and socio-demographic factors associated with visual outcomes post-IOFB removal.

Our main findings were: 1) mean incidence rates of 2.28 per 100,000 patient-years across ophthalmology practices participating in the IRIS Registry, which represent approximately 70% of practicing ophthalmologists across the US^12^; 2) presence of endophthalmitis or RD at baseline was consistently associated with poorer visual outcomes; 3) mean and median VA typically improve following IOFB removal, with a rapid period of recovery by two months that subsequently plateaus; and 4) pre-IOFB VA was the strongest predictor of post-IOFB removal visual outcomes in the subgroup of patients where this was available, with poor VA (logMAR 1.0 or worse) attenuating any improvement in VA.

We found that IOFB injuries disproportionately affect middle-aged males, which may reflect the occupational risk in male-dominated manual trades (e.g. construction, metalwork). The sex predilection was consistent with previous studies in the literature, albeit with lower proportions of males affected compared to the literature (70.4% versus 80-100%).^1–9^ The median age (55 years) in our cohort remained within the working age population, but was one to two decades older than that described in these previous studies, many of which originate from low- and middle-income countries (LMICs), where injuries may more commonly affect younger men engaged in high-risk industrial or agricultural work. The older age and sex distribution may also reflect demographic shifts in the US workforce or the exposure of older individuals to home-based DIY (do-it-yourself) activities. These findings underscore the importance of tailoring injury prevention strategies to evolving risk profiles in high-income countries.

Notably, patients self-identifying as Black or African American had a significantly worse mean visual outcome after controlling for pre-injury VA and a wide range of sociodemographic and clinical presentation factors. Our findings align with reports of racial and ethnic disparities in OGI risk and poorer visual outcomes in the US,^12^ as well as a broader body of evidence documenting disparities in ophthalmic care outcomes for conditions such as RD and glaucoma,^13–15^ and broader systemic trauma outcomes.^16^ However, this disparity may still reflect some degree of unmeasured confounding from social determinants of health, differential access or adherence to follow-up care, or systemic biases within the healthcare system. Given that the association was not observed in a sensitivity analysis using a median-focused mixed effects quantile regression model, the effect of race and ethnicity on visual outcome appears to be driven by a subgroup with particularly poor outcomes. Further research is needed to validate, understand, and address the underlying causes of the racial and ethnic disparities identified which are likely to be social determinants of health, ensuring that efforts to optimize visual outcome and prevention strategies are equitable for all patient populations.

Direct comparison of visual outcome predictors with the published literature is complicated by differing follow-up periods, covariate selection, and statistical modelling strategies. We reviewed recent large studies evaluating predictors of post-IOFB VA. One study of 1176 eyes with IOFB in Southwest China identified predictors broadly consistent with ours, including RD, traumatic cataract, endophthalmitis, and posterior segment IOFB, in a multivariable model examining risk factors for VA >20/200 at discharge. However, additional predictors such as wound size (not available in our study) and poor presenting VA were also reported as predictors.^3^ In contrast, another study of 159 eyes from North China found presenting VA, size of IOFB, size of wound, and macular lesions to be the only factors influencing VA post-IOFB removal.^9^ These variations likely also reflect differences in patient populations, case severity, surgical techniques, and timing of outcome assessment. VA assessment methodology may also vary.^17^ Notably, the follow-up period in these studies was not defined. In our study, we were able to characterize the trajectory of visual recovery up to 18 months post-IOFB removal, which provides new insights into the sustained improvements and plateau phases of visual outcomes after IOFB injury, and may be helpful in informing patient expectations.

Our visual outcome analysis also benefited from the complementary use of two distinct modeling strategies. The linear mixed-effects model, which predicts the mean, was sensitive to extreme outcomes. It identified the presence of endophthalmitis, RD, vitreous hemorrhage, and hyphema at baseline as significant predictors of a poorer visual outcome on average. Sensitivity analyses employing quantile regression, which is more robust to outliers and predicts the median (i.e. typical) outcome, found that only RD and hyphema were associated with worse vision. This divergence suggests that while most patients experiencing RD and hyphema experience worse vision, other patients with complications such as endophthalmitis and vitreous hemorrhage may have a more varied effect, creating a ‘high risk, high variability’ profile, wherein some patients experience reasonable recovery, but where a subset suffers catastrophic vision loss, thereby heavily skewing the overall mean.

### Strengths and limitations

Limitations reflect common challenges inherent to research utilizing large administrative EHR databases such as the IRIS Registry, which rely on the completeness and accuracy of routinely collected clinical and billing data.^18–21^ This is exemplified by the coding for IOFB material – although specific CPT Z-codes exist to specify IOFB material type, these codes were so infrequently used (n=15/4784 cases) as to render the variable analytically unusable. In addition, such registries typically do not contain granular clinical data obtainable from manual chart review of free-text letters (e.g. etiology, injury zone, IOFB material), although advances in natural language processing may make large-scale data extraction possible in the future.^22,23^

Beyond data completeness, the potential for missing data and coding inaccuracies poses another challenge in EHR-based research. For example, pre-IOFB VA was a strong predictor of visual recovery, but pre-IOFB VA data was only available for a subset of patients known to an eye care provider. Given the potential systematic bias (e.g. if these patients were more likely to have pre-existing eye conditions), we opted to conduct a subgroup analysis for the subset of patients with pre-IOFB VA. With regard to the risk of coding inaccuracies, we conducted a review of CPT codes contemporaneous to the time of IOFB removal and identified codes for procedures such as aqueous shunt or anterior segment drainage device revision or insertion, goniotomy, trabeculotomy etc. – interventions which would not be typically performed in the context of OGI repair and IOFB removal. To mitigate this, we adopted a pragmatic and stringent approach in defining our cohort, and incorporated exclusion criteria to remove cases with implausible procedural combinations, with the aim of improving specificity in identifying true IOFB injuries.

Overall, as the largest real-world ophthalmic clinical registry, the IRIS Registry has enabled a robust, population-level analysis of IOFB injuries across a diverse range of socio-demographic and practice settings in the US. The cohort size and national scope provide a powerful foundation for studying clinical questions at scale and estimating trends and outcomes, overcoming the principal constraint of prior literature – namely the limited generalizability of small, single-center series. The large sample size and longer follow-up duration in our cohort allowed us to characterize visual recovery trajectories beyond initial discharge, offering valuable insights into both early and sustained improvements post-IOFB removal, which were not captured in earlier studies. This also facilitated robust evaluation of baseline predictors of visual outcomes. Future work will explore the effect of subsequent complications and procedures.

In summary, this comprehensive longitudinal analysis provides a robust, data-driven model for predicting visual outcomes after IOFB injury and removal. We have mapped the standard VA trajectory experienced post-IOFB removal in a large cohort of patients, demonstrated the importance of pre-IOFB VA in predicting visual outcomes, and quantified the additional risk conferred by specific complications and sociodemographic factors. Together, these findings offer clinicians a model for prognostication to help manage patient expectations, guide clinical decision-making, and ultimately optimize patient care. Future work should aim to integrate granular clinical data to refine these predictive models and facilitate external validation.

## Data Availability

All data referenced in this manuscript are derived from the American Academy of Ophthalmology IRIS® Registry. The Registry is a de-identified, HIPAA-compliant clinical database comprising tens of millions of patient encounters and accessible to researchers via AAO-approved pathways.

## Notes

**Financial support:** This study was funded by the Massachusetts Eye and Ear Clinical Data Science Fund.

**Conflicts of Interest:** None relevant to this work. AYO, EAG, WCK, CR, CA, DAM, SKW, RRS, IL do not have any conflicts to declare. PAK: Cofounder of Cascader Ltd. and has acted as a consultant for Retina Consultants of America, Roche, Boehringer Ingelheim, and Bitfount, and is an equity owner in Big Picture Medical. He has received speaker fees from Zeiss, Thea, Apellis, and Roche. He has received travel support from Bayer and Roche. He has attended advisory boards for Topcon, Bayer, Boehringer-Ingelheim, and Roche. TE: Grant support – Genentech Inc. JWM: Grant support – National Eye Institute, Massachusetts Eye and Ear Clinical Data Science Fund, Lowy Medical Research Institute; Consultant – Genentech/Roche, Heidelberg Engineering, KalVista Pharmaceuticals, ONL Therapeutics, Sunovion; Research support – Aptinyx, Inc., Heidelberg Engineering, KalVista Pharmaceuticals, ONL Therapeutics, Sunovion, Valeant Pharmaceuticals/Mass. Eye and Ear; Patents – ONL Therapeutics, Valeant Pharmaceuticals/Mass. Eye and Ear; Stock – Aptinyx, Inc. AL: Grant support – Massachusetts Eye and Ear Clinical Data Science Fund.

### Competing Interest Statement

AYO, EAG, WCK, CR, CA, DAM, SKW, RRS, IL do not have any conflicts to declare.
PAK: Cofounder of Cascader Ltd. and has acted as a consultant for Retina Consultants of America, Roche, Boehringer Ingelheim, and Bitfount, and is an equity owner in Big Picture Medical. He has received speaker fees from Zeiss, Thea, Apellis, and Roche. He has received travel support from Bayer and Roche. He has attended advisory boards for Topcon, Bayer, Boehringer-Ingelheim, and Roche.
TE: Grant support from Genentech Inc.
JWM: Grant support from National Eye Institute, Massachusetts Eye and Ear Clinical Data Science Fund, Lowy Medical Research Institute; Consultant for Genentech/Roche, Heidelberg Engineering, KalVista Pharmaceuticals, ONL Therapeutics, Sunovion; Research support from Aptinyx, Inc., Heidelberg Engineering, KalVista Pharmaceuticals, ONL Therapeutics, Sunovion, Valeant Pharmaceuticals/Mass. Eye and Ear; Patents for ONL Therapeutics, Valeant Pharmaceuticals/Mass. Eye and Ear; Stock for Aptinyx, Inc.
AL: Grant support from Massachusetts Eye and Ear Clinical Data Science Fund.

### Funding Statement

This study was funded by the Massachusetts Eye and Ear Clinical Data Science Fund.

### Author Declarations

MEE Ophthalmology From: Partners Human Research 399 Revolution Drive, Suite 710 Somerville, MA 02145 Title of Protocol: IRIS Registry Database Project IRB Review Type: Expedited IRB Review Date 02/12/2020 IRB Review Action: Not Human Subjects Research The IRB has determined that this activity does not meet the definition of human subjects research. This determination was based on the PHRC Policy Definition of Human-Subjects Research. Continuing review is not required. The investigators conducting this research will not obtain data through an intervention or interaction with individual subjects or identifiable private information about living individuals. Identifiable private information means that the identity of the individual is or may readily be ascertained by the investigators conducting the research, or is associated with the information. Note: If you are receiving coded data/specimens, please obtain a signed letter of agreement from the provider of the data/specimens stating that you will never be given access to the key to the codes that contains identifiers that could be used to link the samples/data to individual subjects who provided the samples/data. The IRB does not need a copy of this letter of agreement. GENERAL REVIEW COMMENTS: The IRB reviewed and determined that this project does not meet the definition of Human subject Research. Partners Human Research Partners HealthCare 399 Revolution Drive, Suite 710 Somerville, MA 02145 Tel: 857-282-1900 Fax: 857-282-5693

## References

1. Bourke L, Bourke E, Cullinane A, O’Connell E, Idrees Z. Clinical outcomes and epidemiology of intraocular foreign body injuries in Cork University Hospital, Ireland: an 11-year review. Ir J Med Sci. 2021;190(3):1225–1230. doi:10.1007/s11845-020-02443-9

2. Sharma S, Thapa R, Bajimaya S, Pradhan E, Poudyal G. Clinical characteristics and visual outcome, prognostic factor, visual acuity and globe survival in posterior segment intraocular foreign body at Tilganga Institute of Ophthalmology. Nepal J Ophthalmol. 2018;10(19):66–72. doi:10.3126/nepjoph.v10i1.21691

3. Chang T, Zhang Y, Liu L, et al. Epidemiology, Clinical Characteristics, and Visual Outcomes of Patients with Intraocular Foreign Bodies in Southwest China: A 10-Year Review. Ophthalmic Res. 2021;64(3):494–502. doi:10.1159/000513043

4. Santamaría A, Pérez S, De Luis B, Orive A, Feijóo R, Etxebarria J. Clinical characteristics and prognostic factors of open globe injuries in a North Spain population: a 10-year review. Eye. 2023;37(10):2101–2108. doi:10.1038/s41433-022-02297-8

5. Yang Y, Yang C, Zhao R, et al. Intraocular foreign body injury in children: clinical characteristics and factors associated with endophthalmitis. Br J Ophthalmol. 2020;104(6):780–784. doi:10.1136/bjophthalmol-2019-314913

6. Jabłoński M, Winiarczyk M, Biela K, et al. Open Globe Injury (OGI) with a Presence of an Intraocular Foreign Body (IOFB)-Epidemiology, Management, and Risk Factors in Long Term Follow-Up. J Clin Med. 2022;12(1):190. doi:10.3390/jcm12010190

7. Hapca MC, Muntean GA, Drăgan IAN, Vesa LJtefan C, Nicoară SD. Outcomes and Prognostic Factors Following Pars Plana Vitrectomy for Intraocular Foreign Bodies-11-Year Retrospective Analysis in a Tertiary Care Center. J Clin Med. 2022;11(15):4482. doi:10.3390/jcm11154482

8. Isik P, Sizmaz S, Esen E, et al. Management and Clinical Outcomes of Eyes With Posterior Segment Intraocular Foreign Bodies Seen at a Tertiary Referral Center. Ophthalmic Surg Lasers Imaging Retina. 2024;55(8):434–442. doi:10.3928/23258160-20240402-01

9. Xing X, Liu F, Qi Y, Li J, Yu B, Wan L. Clinical Characteristics and Prognostic Factors of Patients with Intraocular Foreign Bodies from a Tertiary Eye Center in North China. Clin Ophthalmol. 2024;18:3635–3643. doi:10.2147/OPTH.S492986

10. McCrum ML, Zakrison TL, Knowlton LM, et al. Taking action to achieve health equity and eliminate healthcare disparities within acute care surgery. Trauma Surg Acute Care Open. 2024;9(1). doi:10.1136/tsaco-2024-001494

11. Chiang MF, Sommer A, Rich WL, Lum F, Parke DW. The 2016 American Academy of Ophthalmology IRIS® Registry (Intelligent Research in Sight) Database. Ophthalmology. 2018;125(8):1143–1148. doi:10.1016/j.ophtha.2017.12.001

12. Tomaiuolo M, Woreta FA, Li A, et al. Open-Globe Injury Repairs in the American Academy of Ophthalmology IRIS® Registry 2014 – 2018. Ophthalmology. 2023;130(8):812–821. doi:10.1016/j.ophtha.2023.03.002

13. Xu J, Davoudi S, Yoon J, et al. Effect of race and ethnicity on surgical outcomes for rhegmatogenous retinal detachments. Canadian Journal of Ophthalmology. 2024;59(2):102–108. doi:10.1016/j.jcjo.2022.12.003

14. Mastropasqua R, Luo YHL, Cheah YS, Egan C, Lewis JJ, da Cruz L. Black patients sustain vision loss while White and South Asian patients gain vision following delamination or segmentation surgery for tractional complications associated with proliferative diabetic retinopathy. Eye. 2017;31(10):1468–1474. doi:10.1038/eye.2017.95

15. Ciociola EC, Sekimitsu S, Smith S, et al. Racial Disparities in Glaucoma Vision Outcomes and Eye Care Utilization: An IRIS Registry Analysis. Am J Ophthalmol. 2024;264:194–204. doi:10.1016/j.ajo.2024.03.022

16. Haider AH, Weygandt PL, Bentley JM, et al. Disparities in trauma care and outcomes in the United States: A systematic review and meta-analysis. J Trauma Acute Care Surg. 2013;74(5):1195–1205. doi:10.1097/TA.0b013e31828c331d

17. Elliott DB. The good (logMAR), the bad (Snellen) and the ugly (BCVA, number of letters read) of visual acuity measurement. Ophthalmic Physiologic Optic. 2016;36(4):355–358. doi:10.1111/opo.12310

18. Kim MK, Rouphael C, McMichael J, Welch N, Dasarathy S. Challenges in and Opportunities for Electronic Health Record-Based Data Analysis and Interpretation. Gut Liver. 2024;18(2):201–208. doi:10.5009/gnl230272

19. Lee CS, Blazes M, Lorch A, et al. American Academy of Ophthalmology Intelligent Research in Sight (IRIS®) Registry and the IRIS Registry Analytic Center Consortium. Ophthalmol Sci. 2022;2(1):100112. doi:10.1016/j.xops.2022.100112

20. Goldberg E, Douglas V, Ivanov A, et al. Data Duplication and Errors in Large Medical Datasets: A Case Study in the IRIS® Registry. Published online March 10, 2025. doi:10.31219/osf.io/gcqkt_v3

21. Ross C, Ivanov A, Elze T, et al. Factors Associated with Missing Sociodemographic Data in the IRIS® (Intelligent Research in Sight) Registry. Ophthalmology Science. 2024;4(6). doi:10.1016/j.xops.2024.100542

22. Stein JD, Rahman M, Andrews C, et al. Evaluation of an Algorithm for Identifying Ocular Conditions in Electronic Health Record Data. JAMA Ophthalmol. 2019;137(5):491–497. doi:10.1001/jamaophthalmol.2018.7051

23. Maganti N, Tan H, Niziol LM, et al. Natural Language Processing to Quantify Microbial Keratitis Measurements. Ophthalmology. 2019;126(12):1722–1724. doi:10.1016/j.ophtha.2019.06.003

