## Supplementary material for "Epidemiology, clinical features, and visual outcomes after intraocular foreign body removal: an IRIS® Registry (Intelligent Research in Sight) Analysis": Table 1

**Table 1: Frequency and incidence of IOFB cases undergoing surgery in the IRIS Registry.**

| **Year** | **IOFB (N)*** | **Total population in that year (N)** | **Incidence with 95% CI per 100,000 patient-years** |
| --- | --- | --- | --- |
| **2016** | 456 | 17,518,467 | 2.60 (2.37, 2.85) |
| **2017** | 546 | 19,232,704 | 2.84 (2.61, 3.10) |
| **2018** | 573 | 20,654,335 | 2.77 (2.55, 3.01) |
| **2019** | 532 | 22,258,778 | 2.39 (2.19, 2.60) |
| **2020** | 541 | 21,137,177 | 2.56 (2.35, 2.79) |
| **2021** | 600 | 25,515,248 | 2.35 (2.17, 2.55) |
| **2022** | 550 | 27,233,664 | 2.02 (1.85, 2.20) |
| **2023** | 573 | 29,324,229 | 1.95 (1.80, 2.12) |
| **2024^** | 346 | 23,820,914 | 1.45 (1.30, 1.61) |

Legend: CI- confidence interval; IOFB- intraocular foreign body; N- number.

*Bilateral IOFB cases undergoing surgery on the same date were treated as 1 case (patient-level analysis for incidence calculations).

^Data for 2024 were only available till October 2024.
