## Supplementary material for "Epidemiology, clinical features, and visual outcomes after intraocular foreign body removal: an IRIS® Registry (Intelligent Research in Sight) Analysis": Table 2

**Table 2: Socio-demographic characteristics of the overall patient cohort.**

|  |  | **N (%)** |
| --- | --- | --- |
| **Age (Median, IQR) (years)** |  | 55 (36-70) |
| **Sex** | **Female** | 1311 (27.4) |
|  | **Male** | 3362 (70.3) |
|  | **Unknown** | 111 (2.3) |
| **High school graduation percentage** | **≤ 60** | 28 (0.6) |
|  | **61-70** | 127 (2.7) |
|  | **71-80** | 414 (8.7) |
|  | **81-90** | 1,435 (30.0) |
|  | **91-100** | 2,117 (44.3) |
|  | **Unknown** | 663 (13.9) |
| **Urban/ Rural Status** | **Urban** | 3950 (82.6) |
|  | **Rural** | 799 (16.7) |
|  | **Unknown** | 35 (0.7) |
| **Median Income** | **≤$34,999** | 138 (2.9) |
|  | **$35,000 - $74,999** | 2,488 (52.0) |
|  | **$75,000 - $149,999** | 1,375 (28.7) |
|  | **≥ $150,000** | 106 (2.2) |
|  | **Unknown** | 677 (14.2) |
| **Race** | **White** | 2,853 (59.6) |
|  | **Asian** | 96 (2.0) |
|  | **Black or African American** | 382 (8.0) |
|  | **Other** | 590 (12.3) |
|  | **Unknown** | 863 (18.0) |
| **Ethnicity** | **Hispanic or Latino** | 604 (12.6) |
|  | **Not Hispanic or Latino** | 2875 (60.1) |
|  | **Unknown** | 1305 (27.3) |
| **Practice Region** | **Midwest** | 697 (14.6) |
|  | **Northeast** | 703 (14.7) |
|  | **South** | 2,039 (42.6) |
|  | **West** | 854 (17.9) |
|  | **US Territory** | 13 (0.3) |
|  | **Unknown** | 478 (10.0) |

Legend: IQR, interquartile range; N, number.
