## Supplementary material for "Epidemiology, clinical features, and visual outcomes after intraocular foreign body removal: an IRIS® Registry (Intelligent Research in Sight) Analysis": Table 3

**Table 3: Clinical characteristics and procedures performed at baseline.**

|  |  | **N (%)** |
| --- | --- | --- |
| **Unilateral** |  | 4636 (96.9) |
| **IOFB Location** | **Anterior segment** | 2229 (46.6) |
|  | **Posterior segment** | 2478 (51.8) |
|  | **Both** | 77 (1.6) |
| **IOFB Extraction Method** | **Not recorded** | 2229 (46.6) |
|  | **Non-magnetic** | 1958 (40.9) |
|  | **Magnetic** | 581 (12.1) |
|  | **Both** | 16 (0.3) |
| **Concurrent Procedures** | **PPV** | 2032 (42.5) |
|  | **Repair of cornea and/or sclera and/or anterior segment laceration** | 853 (17.8) |
|  | **AC washout** | 700 (14.6) |
|  | **Lens extraction** | 548 (11.5) |
|  | **Intravitreal injection (drug not specified)** | 179 (3.7) |
|  | **AC paracentesis** | 139 (2.9) |
|  | **Intravitreal injection (Ceftazidime/ Vancomycin)** | 86 (1.8) |
|  | **Anterior vitrectomy** | 42 (0.9) |
| **Ocular Investigations** | **B-scan** | 432 (9.0) |

Legend: IOFB, intraocular foreign body; AC, anterior chamber; PPV, pars plana vitrectomy; N, number.
