## Supplementary material for "Epidemiology, clinical features, and visual outcomes after intraocular foreign body removal: an IRIS® Registry (Intelligent Research in Sight) Analysis": Table 4

**Table 4: Incident complications at baseline, early (1-6 days), and late (7 days to 18 months).**

|  | **Baseline (N, %)** | **Early (N, %)** | **Late (N, %)** |
| --- | --- | --- | --- |
| **Retinal detachment** | 597 (12.5) | 52 (1.1) | 633 (13.2) |
| **Cataract** | 501 (10.5) | 78 (1.6) | 749 (15.7) |
| **Vitreous hemorrhage** | 378 (7.9) | 74 (1.5) | 153 (3.2) |
| **Endophthalmitis** | 187 (3.9) | 12 (1.7) | 27 (0.6) |
| **Hyphema** | 160 (3.3) | 36 (0.8) | 67 (1.4) |
| **Retinal tear** | 130 (2.7) | 27 (0.6) | 93 (1.9) |
| **Raised intraocular pressure or glaucoma** | 86 (1.7) | 36 (0.8) | 354 (7.4) |
| **Proliferative vitreoretinopathy** | 21 (0.4) | 7 (0.2) | 92 (1.9) |
| **Iridodialysis** | 6 (0.1) | 1 (0.0) | 10 (0.2) |
| **Enucleation** |  | 2 (0.0) | 17 (0.4) |
| **Evisceration** |  | 2 (0.0) | 9 (0.2) |
| **Corneal scar** |  |  | 237 (5.0) |
| **Siderosis** |  |  | 4 (0.1) |
| **Traumatic optic neuropathy** |  |  | 3 (0.1) |
| **Sympathetic ophthalmia** |  |  | 1 (0.0) |

Legend: N, number.
