## Supplementary S1 for "Epidemiology, clinical features, and visual outcomes after intraocular foreign body removal: an IRIS® Registry (Intelligent Research in Sight) Analysis"

**Supplementary S1: International Classification of Diseases 10^th^ Edition (ICD-10), Systematized Nomenclature of Medicine Clinical Terms (SNOMED-CT), and Current Procedural Terminology (CPT) codes used to define the eligibility criteria for the study cohort.**

**Inclusion criteria:**

CPT codes addressing the removal of IOFB (first-ever instance):

- 65235 Removal of foreign body, intraocular; from anterior chamber of eye or lens
- 65260 Removal of foreign body, intraocular; from posterior segment, magnetic extraction, anterior or posterior route
- 65265 Removal of foreign body, intraocular; from posterior segment, nonmagnetic extraction

**To which the following exclusion criteria were applied:**

ICD-10 codes:

- H59.021 Cataract (lens) fragments in eye following cataract surgery, right eye
- H59.022 Cataract (lens) fragments in eye following cataract surgery, left eye
- H59.023 Cataract (lens) fragments in eye following cataract surgery, bilateral

CPT codes

- 67121 Removal of implanted material, posterior segment; intraocular
- 66985 Insertion of intraocular lens prosthesis (secondary implant), not associated with concurrent cataract removal
- 66986 Exchange of intraocular lens
- 66825 Repositioning of intraocular lens prosthesis, requiring an incision (separate procedure)
- 65920 Removal of implanted material, anterior segment of eye
- 67120 Removal of implanted material, posterior segment; extraocular
- 66180 Aqueous shunt to extraocular equatorial plate reservoir, external approach; with graft
- 66185 Revision of aqueous shunt to extraocular equatorial plate reservoir; with graft
- 0191T Insertion of anterior segment aqueous drainage device, without extraocular reservoir, internal approach, into the trabecular meshwork; initial insertion
- 66183 Insertion of anterior segment aqueous drainage device, without extraocular reservoir, external approach
- 66174 Transluminal dilation of aqueous outflow canal (e.g., canaloplasty); without retention of device or stent
- 0376T Insertion of anterior segment aqueous drainage device, without extraocular reservoir, internal approach, into the trabecular meshwork; each additional device insertion (List separately in addition to code for primary procedure)
- 0449T Insertion of aqueous drainage device, without extraocular reservoir, internal approach, into the subconjunctival space; initial device
- 0671T Insertion of anterior segment aqueous drainage device into the trabecular meshwork, without external reservoir, and without concomitant cataract removal, one or more
- 66179 Aqueous shunt to extraocular equatorial plate reservoir, external approach; without graft
- 0253T Insertion of anterior segment aqueous drainage device, without extraocular reservoir, internal approach, into the suprachoroidal space
- 65850 Trabeculotomy ab externo
- 65820 Goniotomy
- 66170 Fistulization of sclera for glaucoma; trabeculectomy ab externo in absence of previous surgery
- 66172 Fistulization of sclera for glaucoma; trabeculectomy ab externo with scarring from previous ocular surgery or trauma (includes injection of antifibrotic agents)
- 66184 Revision of aqueous shunt to extraocular equatorial plate reservoir; without graft
- 65865 Severing adhesions of anterior segment of eye, incisional technique (with or without injection of air or liquid) (separate procedure); goniosynechiae
