## Supplementary Table 2 for "Epidemiology, clinical features, and visual outcomes after intraocular foreign body removal: an IRIS® Registry (Intelligent Research in Sight) Analysis"

**Supplementary Table 2: ICD-10 codes used to extract intraocular foreign body-associated complications at baseline; these complications were identified from a literature review.**

| **Complication** | **ICD-10 code** |
| --- | --- |
| Endophthalmitis | H44.0 Post-traumatic endophthalmitis  Condition_code like 'H44.00%'  or condition_code like 'H44.19%' |
| Glaucoma or raised intraocular pressure | H40.3 Glaucoma secondary to eye trauma  H40.8 Other glaucoma  H40.9 Glaucoma, unspecified  H40.05 Increased intraocular pressure |
| Proliferative vitreoretinopathy | H44.13 Proliferative vitreoretinopathy |
| Cataract | H26.1: Traumatic cataract  H26.9: Unspecified cataract  H25.20: Age-related cataract  H25.89: Other age-related cataract  H25.10: Age-related nuclear cataract  H26.2: Complicated cataract  H26.4: Secondary cataract  H26.8: Other specified cataract |
| Hyphema | H21.0 Hyphema |
| Vitreous hemorrhage | H43.1 Vitreous hemorrhage |
| Iridodialysis | H21.539 Iridodialysis |
| Retinal tear | H33.3 Retinal tear |
| Retinal detachment | H33.20 Retinal detachment  H33.033 Retinal detachment of both eyes with giant retinal tear  H33.023 Retinal detachment of both eyes with multiple breaks  H33.23 Retinal detachment of both eyes with presence of subretinal fluid  H33.029 Retinal detachment with multiple breaks, unspecified eye  H33.003 Retinal detachment of both eyes with retinal break  H33.043 Retinal detachment of both eyes with retinal dialysis  H33.013 Retinal detachment of both eyes with single break  H33.001 Unspecified retinal detachment with retinal break, right eye  H33.011 Retinal detachment with single break, right eye  H33.021 Retinal detachment with multiple breaks, right eye  H33.21 Retinal detachment of right eye with presence of subretinal fluid  H33.031 Retinal detachment with giant retinal tear, right eye  H33.041 Retinal detachment with retinal dialysis, right eye  H33.051 Total retinal detachment, right eye  H33.41 Traction detachment of retina, right eye  H33.002 Unspecified retinal detachment with retinal break, left eye  H33.012 Retinal detachment with single break, left eye  H33.022 Retinal detachment with multiple breaks, left eye  H33.22 Retinal detachment of left eye with presence of subretinal fluid  H33.032 Retinal detachment with giant retinal tear, left eye  H33.042 Retinal detachment with retinal dialysis, left eye  H33.052 Total retinal detachment, left eye  H33.009 Retinal detachment with break  H33.039 Retinal detachment with giant retinal tear, unspecified eye  H33.40 Traction detachment of retina, unspecified eye  H33.41 Tractional detachment of retina, right eye  H33.42 Traction detachment of retina, left eye  H33.8 Other retinal detachments  H33.43 Traction detachment of retina, bilateral  H33.009 Retinal Detachment with break  H33.039 Retinal detachment with giant retinal tear, unspecified eye |

Legend: ICD-10, International Classification of Diseases 10th Revision
