## Supplementary Table 3 for "Epidemiology, clinical features, and visual outcomes after intraocular foreign body removal: an IRIS® Registry (Intelligent Research in Sight) Analysis"

**Supplementary Table 3: Socio-demographic characteristics of the overall patient cohort, and in each year from 2016-2024. Note that data for 2024 is only available untill October 2024.**

|  |  | **Overall (N,%)** | **2016 (N,%)** | **2017 (N,%)** | **2018 (N,%)** | **2019 (N,%)** | **2020 (N,%)** | **2021 (N,%)** | **2022 (N,%)** | **2023 (N,%)** | **2024 (N,%)** |
| --- | --- | --- | --- | --- | --- | --- | --- | --- | --- | --- | --- |
| **Age (Median, IQR) (years)** | | 55 (36-70) | 58 (36-72) | 56 (38-70) | 56 (38-71) | 54 (34-69) | 54 (35-67) | 55 (35-70) | 53 (34-69) | 54 (36-70) | 60 (41-72) |
| **Sex** | **Female** | 1311 (27.4) | 137 (29.4) | 151 (27.0) | 154 (26.4) | 161 (29.9) | 151 (27.7) | 157 (25.8) | 136 (24.5) | 154 (26.6) | 110 (31.5) |
|  | **Male** | 3362 (70.3) | 318 (68.2) | 394 (70.5) | 405 (69.5) | 368 (68.4) | 381 (69.8) | 445 (73.1) | 405 (72.8) | 412 (71.3) | 234 (67.0) |
|  | **Unknown** | 111 (2.3) | 11 (2.4) | 14 (2.5) | 24 (4.1) | 9 (1.7) | 14 (2.6) | 7 (1.1) | 15 (2.7) | 12 (2.1) | 5 (1.4) |
| **High school graduation percentage** | **≤ 60** | 28 (0.6) | 2 (0.4) | 3 (0.5) | 2 (0.3) | 3 (0.6) | 2 (0.4) | 5 (0.8) | 4 (0.7) | 2 (0.3) | 5 (1.4) |
|  | **61-70** | 127 (2.7) | 2 (0.4) | 3 (0.5) | 2 (0.3) | 3 (0.6) | 2 (0.4) | 5 (0.8) | 4 (0.7) | 2 (0.3) | 5 (1.4) |
|  | **71-80** | 414 (8.7) | 13 (2.8) | 6 (1.1) | 15 (2.6) | 12 (2.2) | 16 (2.9) | 16 (2.6) | 20 (3.6) | 20 (3.5) | 9 (2.6) |
|  | **81-90** | 1,435 (30.0) | 40 (8.6) | 42 (7.5) | 62 (10.6) | 48 (8.9) | 46 (8.4) | 48 (7.9) | 46 (8.3) | 55 (9.5) | 27 (7.7) |
|  | **91-100** | 2,117 (44.3) | 136 (29.2) | 168 (30.1) | 161 (27.6) | 147 (27.3) | 185 (33.9) | 191 (31.4) | 170 (30.6) | 186 (32.2) | 91 (26.1) |
|  | **Unknown** | 663 (13.9) | 227 (48.7) | 275 (49.2) | 271 (46.5) | 240 (44.6) | 219 (40.1) | 262 (43.0) | 226 (40.6) | 237 (41.0) | 160 (45.8) |
| **Urban/ Rural Status** | **Urban** | 3950 (82.6) | 388 (83.3) | 459 (82.1) | 489 (83.9) | 450 (83.6) | 453 (83.0) | 509 (83.6) | 451 (81.1) | 467 (80.8) | 284 (81.4) |
|  | **Rural** | 799 (16.7) | 77 (16.5) | 91 (16.3) | 91 (15.6) | 83 (15.4) | 91 (16.7) | 95 (15.6) | 103 (18.5) | 107 (18.5) | 16 (17.5) |
|  | **Unknown** | 35 (0.7) | 1 (0.2) | 9 (1.6) | 3 (0.5) | 5 (0.9) | 2 (0.4) | 5 (0.8) | 2 (0.4) | 4 (0.7) | 4 (1.1) |
| **Median Income** | **≤$34,999** | 138 (2.9) | 16 (3.4) | 19 (3.4) | 14 (2.4) | 12 (2.2) | 15 (2.7) | 20 (3.3) | 15 (2.7) | 13 (2.2) | 14 (4.0) |
|  | **$35,000 - $74,999** | 2,488 (52.0) | 252 (54.1) | 289 (51.7) | 314 (53.9) | 271 (50.4) | 292 (53.5) | 305 (50.1) | 294 (52.9) | 312 (54.0) | 159 (45.6) |
|  | **$75,000 - $149,999** | 1,375 (28.7) | 145 (31.1) | 171 (30.6) | 159 (27.3) | 153 (28.4) | 150 (27.5) | 180 (29.6) | 145 (26.1) | 163 (28.2) | 109 (31.2) |
|  | **≥$150,000** | 106 (2.2) | 4 (0.9) | 14 (2.5) | 23 (3.9) | 13 (2.4) | 8 (1.5) | 16 (2.6) | 11 (2.0) | 9 (1.6) | 8 (2.3) |
|  | **Unknown** | 677 (14.2) | 49 (10.5) | 66 (11.8) | 73 (12.5) | 89 (16.5) | 81 (14.8) | 88 (14.4) | 91 (16.4) | 81 (14.0) | 59 (16.9) |
| **Race** | **White** | 2,853 (59.6) | 296 (63.5) | 361 (64.6) | 359 (61.6) | 315 (58.6) | 338 (61.9) | 349 (57.3) | 313 (56.3) | 336 (58.1) | 186 (53.3) |
|  | **Asian** | 96 (2.0) | 8 (1.7) | 9 (1.6) | 15 (2.6) | 19 (3.5) | 10 (1.8) | 11 (1.8) | 9 (1.6) | 8 (1.4) | 7 (2.0) |
|  | **Black or African American** | 382 (8.0) | 26 (5.6) | 58 (10.4) | 41 (7.0) | 36 (6.7) | 56 (10.3) | 56 (9.2) | 37 (6.7) | 43 (7.4) | 29 (8.3) |
|  | **Other** | 590 (12.3) | 43 (9.2) | 53 (9.5) | 63 (10.8) | 77 (14.3) | 70 (12.8) | 80 (13.1) | 86 (15.5) | 70 (12.1) | 48 (13.8) |
|  | **Unknown** | 863 (18.0) | 93 (20.0) | 78 (14.0) | 105 (18.0) | 91 (16.9) | 72 (13.2) | 113 (18.6) | 111 (20.0) | 121 (20.9) | 79 (22.6) |
| **Ethnicity** | **Hispanic or Latino** | 604 (12.6) | 66 (14.2) | 63 (11.3) | 68 (11.7) | 76 (14.1) | 72 (13.2) | 75 (12.3) | 70 (12.6) | 66 (11.4) | 48 (13.8) |
|  | **Not Hispanic or Latino** | 2875 (60.1) | 312 (67.0) | 397 (71.0) | 380 (65.2) | 309 (57.4) | 335 (61.4) | 343 (56.3) | 303 (54.5) | 314 (54.3) | 182 (52.1) |
|  | **Unknown** | 1305 (27.3) | 88 (18.9) | 99 (17.7) | 135 (23.2) | 153 (28.4) | 139 (25.5) | 191 (31.4) | 183 (32.9) | 198 (34.3) | 119 (34.1) |
| **Practice Region** | **Midwest** | 697 (14.6) | 78 (16.7) | 77 (13.8) | 84 (14.4) | 58 (10.8) | 74 (13.6) | 96 (15.8) | 80 (14.4) | 89 (15.4) | 61 (17.5) |
|  | **Northeast** | 703 (14.7) | 60 (12.9) | 65 (11.6) | 81 (13.9) | 85 (15.8) | 78 (14.3) | 116 (19.0) | 70 (12.6) | 90 (15.6) | 58 (16.6) |
|  | **South** | 2,039 (42.6) | 187 (40.1) | 228 (40.8) | 230 (39.5) | 217 (40.3) | 275 (50.4) | 254 (41.7) | 265 (47.7) | 251 (43.4) | 132 (37.8) |
|  | **Unknown** | 478 (10.0) | 46 (9.9) | 74 (13.2) | 75 (12.9) | 59 (11.0) | 45 (8.2) | 55 (9.0) | 48 (8.6) | 52 (9.0) | 24 (6.9) |
|  | **US Territory** | 13 (0.3) | 1 (0.2) | 1 (0.2) | 3 (0.5) | 3 (0.6) | 1 (0.2) | 0 (0.0) | 1 (0.2) | 1 (0.2) | 2 (0.6) |
|  | **West** | 854 (17.9) | 94 (20.2) | 114 (20.4) | 110 (18.9) | 116 (21.6) | 73 (13.4) | 88 (14.4) | 92 (16.5) | 95 (16.4) | 72 (20.6) |

Legend: IQR, interquartile range; N, number
