## Supplementary Table 4 for "Epidemiology, clinical features, and visual outcomes after intraocular foreign body removal: an IRIS® Registry (Intelligent Research in Sight) Analysis"

**Supplementary Table 4: Mixed-effects model for predicting change in visual acuity relative to baseline post-IOFB removal.**

| **Predictor (Fixed effects)** | **Estimate (β)** | **95% CI** | **p-value** |
| --- | --- | --- | --- |
| (Intercept) | 0.99 | 0.88 – 1.10 | <0.001 |
| Time period (ref: Baseline) |  |  |  |
| 1-month post-IOFB | 0.01 | -0.02 – 0.04 | 0.637 |
| 2 months post-IOFB | -0.38 | -0.41 – -0.34 | <0.001 |
| 3 months post-IOFB | -0.43 | -0.47 – -0.39 | <0.001 |
| 4 months post-IOFB | -0.43 | -0.47 – -0.39 | <0.001 |
| 5 months post-IOFB | -0.48 | -0.53 – -0.43 | <0.001 |
| 6 months post-IOFB | -0.46 | -0.51 – -0.41 | <0.001 |
| 7 months post-IOFB | -0.49 | -0.54 – -0.43 | <0.001 |
| 8 months post-IOFB | -0.47 | -0.53 – -0.41 | <0.001 |
| 9 months post-IOFB | -0.48 | -0.55 – -0.42 | <0.001 |
| 10 months post-IOFB | -0.50 | -0.57 – -0.44 | <0.001 |
| 11 months post-IOFB | -0.50 | -0.57 – -0.43 | <0.001 |
| 12 months post-IOFB | -0.54 | -0.62 – -0.47 | <0.001 |
| 13 months post-IOFB | -0.53 | -0.61 – -0.45 | <0.001 |
| 14 months post-IOFB | -0.55 | -0.63 – -0.46 | <0.001 |
| 15 months post-IOFB | -0.46 | -0.55 – -0.37 | <0.001 |
| 16 months post-IOFB | -0.57 | -0.67 – -0.48 | <0.001 |
| 17 months post-IOFB | -0.55 | -0.65 – -0.45 | <0.001 |
| 18 months post-IOFB | -0.59 | -0.69 – -0.48 | <0.001 |
| Sociodemographic factors |  |  |  |
| Age (per decade; reference: <65) | 0.02 | -0.01 – 0.04 | 0.229 |
| Sex: Female (ref: Male) | 0.11 | 0.00 – 0.21 | 0.084 |
| Race: Asian (ref: White) | 0.05 | -0.25 – 0.36 | 0.805 |
| Race: Black or African American (ref: White) | 0.35 | 0.17 – 0.52 | <0.001 |
| Race: Other (ref: White) | -0.06 | -0.50 – 0.39 | 0.835 |
| Race: Unknown (ref: White) | -0.07 | -0.20 – 0.05 | 0.376 |
| Ethnicity: Hispanic or Latino (ref: Not Hispanic or Latino) | 0.07 | -0.08 – 0.22 | 0.485 |
| Ethnicity: Unknown (ref: Not Hispanic or Latino) | -0.16 | -0.26 – -0.05 | 0.005 |
| Median income: USD 34,999 and below (ref: 35,000-74,999) | 0.05 | -0.21-0.30 | 0.805 |
| Median income: USD 75,000-149,999 (ref: 35,000-74,999) | -0.06 | -0.16-0.05 | 0.394 |
| Median income: USD 150,000 and above (ref: 35,000-74,999) | -0.15 | -0.48-0.17 | 0.468 |
| Median income: Unknown (ref: 35,000-74,999) | 0.11 | -0.80-1.01 | 0.835 |
| Urban/Rural status: Rural (ref: Urban) | -0.02 | -0.13-0.09 | 0.805 |
| Clinical factors |  |  |  |
| IOFB location: Posterior segment (ref: anterior segment) | 0.25 | 0.15 – 0.35 | <0.001 |
| IOFB location: Both anterior and posterior (ref: anterior segment) | 0.58 | 0.07 – 1.09 | 0.045 |
| Presenting complication: Endophthalmitis | 0.70 | 0.56 – 0.84 | <0.001 |
| Presenting complication: Hyphema | 0.74 | 0.55 – 0.93 | <0.001 |
| Presenting complication: Vitreous hemorrhage | 0.23 | 0.10 – 0.36 | 0.001 |
| Presenting complication: Retinal tear | -0.14 | -0.38 – 0.11 | 0.380 |
| Presenting complication: Retinal detachment | 0.35 | 0.22 – 0.48 | <0.001 |
| Presenting complication: Cataract | 0.13 | 0.02 – 0.24 | 0.029 |

**Legend**: CI, confidence interval; IOFB, intraocular foreign body.

Higher estimates (β) indicate worse visual acuity. Reference groups are baseline (time period 0), absent (for clinical factors), anterior (for IOFB location), White (for race), Not Hispanic or Latino (for ethnicity), and Male (for sex).

Random Effects: Residual variance (σ²): 0.25; Intercept variance (eye ID) (τ₀₀): 0.56; Slope variance (time) (τ₁₁): 0.00; Correlation (intercept–slope) (ρ₀₁): 0.09; Intraclass correlation (ICC): 0.69. Model Fit: Observations: 15,882; Marginal R²: 0.200; Conditional R²: 0.750
