## Supplementary Table 5 for "Epidemiology, clinical features, and visual outcomes after intraocular foreign body removal: an IRIS® Registry (Intelligent Research in Sight) Analysis"

**Supplementary Table 5: Sensitivity analysis using a mixed-effects quantile regression model for median visual acuity.**

| **Predictor (Fixed effects)** | **Estimate (β)** | **Std Error** | **95% CI** | **Adjusted p-value** |
| --- | --- | --- | --- | --- |
| (Intercept) | 1.089 | 0.109 | 0.871 – 1.308 |  |
| Time period (ref: Baseline) |  |  |  |  |
| 1-month post-IOFB | 0.080 | 0.032 | 0.02 – 0.15 | 0.033 |
| 2 months post-IOFB | -0.213 | 0.036 | -0.29 – -0.14 | < 0.001 |
| 3 months post-IOFB | -0.250 | 0.037 | -0.33 – -0.18 | < 0.001 |
| 4 months post-IOFB | -0.260 | 0.042 | -0.34 – -0.18 | < 0.001 |
| 5 months post-IOFB | -0.282 | 0.037 | -0.36 – -0.21 | < 0.001 |
| 6 months post-IOFB | -0.267 | 0.042 | -0.35 – -0.18 | < 0.001 |
| 7 months post-IOFB | -0.260 | 0.046 | -0.35 – -0.17 | < 0.001 |
| 8 months post-IOFB | -0.276 | 0.042 | -0.36 – -0.19 | < 0.001 |
| 9 months post-IOFB | -0.287 | 0.042 | -0.37 – -0.20 | < 0.001 |
| 10 months post-IOFB | -0.302 | 0.045 | -0.39 – -0.21 | < 0.001 |
| 11 months post-IOFB | -0.283 | 0.046 | -0.38 – -0.19 | < 0.001 |
| 12 months post-IOFB | -0.253 | 0.050 | -0.35 – -0.15 | < 0.001 |
| 13 months post-IOFB | -0.272 | 0.044 | -0.37 – -0.17 | < 0.001 |
| 14 months post-IOFB | -0.302 | 0.050 | -0.40 – -0.20 | < 0.001 |
| 15 months post-IOFB | -0.287 | 0.053 | -0.39 – -0.18 | < 0.001 |
| 16 months post-IOFB | -0.321 | 0.053 | -0.43 – -0.21 | < 0.001 |
| 17 months post-IOFB | -0.330 | 0.053 | -0.44 – -0.22 | < 0.001 |
| 18 months post-IOFB | -0.378 | 0.050 | -0.48 – -0.28 | < 0.001 |
| Sociodemographic factors |  |  |  |  |
| Age (per decade; reference: <65) | 0.029 | 0.025 | -0.02 – 0.08 | 0.367 |
| Sex: Female (ref: Male) | -0.048 | 0.092 | -0.23 – 0.14 | 0.672 |
| Race: Asian (ref: White) | 0.227 | 0.323 | -0.42 – 0.88 | 0.570 |
| Race: Black or African American (ref: White) | 0.132 | 0.177 | -0.22 – 0.49 | 0.558 |
| Race: Other (ref: White) | 0.222 | 0.284 | -0.35 – 0.79 | 0.552 |
| Race: Unknown (ref: White) | 0.126 | 0.124 | -0.12 – 0.38 | 0.435 |
| Ethnicity: Hispanic or Latino (ref: Not Hispanic or Latino) | 0.165 | 0.123 | -0.08 – 0.41 | 0.296 |
| Ethnicity: Unknown (ref: Not Hispanic or Latino) | -0.112 | 0.086 | -0.29 – 0.06 | 0.309 |
| Median income: USD 34,999 and below (ref: 35,000-74,999) | 0.144 | 0.212 | -0.28 – 0.57 | 0.573 |
| Median income: USD 75,000-149,999 (ref: 35,000-74,999) | -0.002 | 0.080 | -0.16 – 0.16 | 0.982 |
| Median income: USD 150,000 and above (ref: 35,000-74,999) | -0.179 | 0.230 | -0.64 – 0.28 | 0.552 |
| Median income: Unknown (ref: 35,000-74,999) | 0.023 | 0.250 | -0.48 – 0.53 | 0.974 |
| Urban/Rural status: Rural (ref: Urban) | 0.025 | 0.102 | -0.18 – 0.23 | 0.870 |
| Clinical factors |  |  |  |  |
| IOFB location: Posterior segment (ref: anterior segment) | 0.091 | 0.076 | -0.06 – 0.24 | 0.355 |
| IOFB location: Both anterior and posterior (ref: anterior segment) | 0.499 | 0.215 | -0.07 – 0.93 | 0.047 |
| Presenting complication: Endophthalmitis | 0.420 | 0.183 | 0.05 – 0.79 | 0.047 |
| Presenting complication: Hyphema | 0.300 | 0.194 | -0.09 – 0.69 | 0.214 |
| Presenting complication: Vitreous hemorrhage | 0.225 | 0.107 | 0.01 – 0.44 | 0.071 |
| Presenting complication: Retinal tear | 0.011 | 0.165 | -0.32 – 0.34 | 0.974 |
| Presenting complication: Retinal detachment | 0.418 | 0.126 | 0.17 – 0.67 | 0.004 |
| Presenting complication: Cataract | -0.074 | 0.095 | -0.26 – 0.12 | 0.552 |

**Legend:** CI, confidence interval; IOFB, intraocular foreign body.

Higher estimates (β) indicate worse visual acuity. P-values are adjusted for multiple comparisons using the Benjamini-Hochberg method. Reference groups are baseline (time period 0), absent (for clinical factors), anterior (for IOFB location), White (for race), Not Hispanic or Latino (for ethnicity), and Male (for sex).
