## Supplementary Table 6 for "Epidemiology, clinical features, and visual outcomes after intraocular foreign body removal: an IRIS® Registry (Intelligent Research in Sight) Analysis"

**Supplementary Table 6: Clinical characteristics comparing subgroup eyes with pre-IOFB and post-IOFB VA, versus the wider cohort.**

|  |  | **Wider Cohort (N, %)** | **Subgroup with pre-IOFB VA (N, %)** | **p-value** |
| --- | --- | --- | --- | --- |
| **Age (Median, IQR) (years)** |  | 55 (36-70) | 68 (58-76) | <0.001 |
| **Sex** | **Female** | 1311 (27.4) | 270 (41.2) | <0.001 |
|  | **Male** | 3362 (70.3) | 371 (56.6) |  |
|  | **Unknown** | 111 (2.3) | 15 (2.3) |  |
| **High school graduation percentage** | **≤ 60** | 28 (0.6) | 2 (0.3) | 0.12 |
|  | **61-70** | 127 (2.7) | 16 (2.4) |  |
|  | **71-80** | 414 (8.7) | 44 (6.7) |  |
|  | **81-90** | 1,435 (30.0) | 176 (26.8) |  |
|  | **91-100** | 2,117 (44.3) | 234 (49.4) |  |
|  | **Unknown** | 663 (13.9) | 94 (14.3) |  |
| **Urban/ Rural Status** | **Urban** | 3950 (82.6) | 68 (10.4) | <0.001 |
|  | **Rural** | 799 (16.7) | 587 (89.5) |  |
|  | **Unknown** | 35 (0.7) | 1 (0.2) |  |
| **Median Income** | **<$34,999** | 138 (2.9%) | 19 (2.9) | 0.004 |
|  | **$35,000 - $74,999** | 2,488 (52.0%) | 292 (44.5) |  |
|  | **$75,000 - $149,999** | 1,375 (28.7%) | 229 (34.9) |  |
|  | **> $150,000** | 106 (2.2%) | 19 (2.9) |  |
|  | **Unknown** | 677 (14.2%) | 97 (14.8) |  |
| **Race** | **White** | 2,853 (59.6%) | 409 (62.3) | <0.001 |
|  | **Asian** | 96 (2.0%) | 25 (3.8) |  |
|  | **Black or African American** | 382 (8.0%) | 58 (8.8) |  |
|  | **Other** | 590 (12.3%) | 85 (13.0) |  |
|  | **Unknown** | 863 (18.0%) | 79 (12.0) |  |
| **Ethnicity** | **Hispanic or Latino** | 604 (12.6) | 61 (9.3) | <0.001 |
|  | **Not Hispanic or Latino** | 2875 (60.1) | 457 (69.7) |  |
|  | **Unknown** | 1305 (27.3) | 138 (21.0) |  |
| **Practice Region** | **Midwest** | 697 (14.6%) | 91 (13.9) | <0.001 |
|  | **Northeast** | 703 (14.7%) | 138 (21.0) |  |
|  | **South** | 2,039 (42.6%) | 224 (34.1) |  |
|  | **Unknown** | 478 (10.0%) | 83 (12.7) |  |
|  | **US Territory** | 13 (0.3%) | 3 (0.5) |  |
|  | **West** | 854 (17.9%) | 117 (17.8) |  |
| **IOFB Location** | **Anterior segment** | 2229 (46.6) | 469 (71.5) | <0.001 |
|  | **Posterior segment** | 2478 (51.8) | 182 (27.7) |  |
|  | **Both** | 77 (1.6) | 5 (0.8) |  |
| **Complications at baseline** | **Retinal detachment** | 597 (12.5) | 46 (0.7) | <0.001 |
|  | **Cataract** | 501 (10.5) | 38 (5.8) | <0.001 |
|  | **Vitreous hemorrhage** | 378 (7.9) | 33 (5.0) | 0.009 |
|  | **Endophthalmitis** | 187 (3.9) | 62 (9.5) | <0.001 |
|  | **Hyphema** | 160 (3.3) | 51 (7.8) | <0.001 |
|  | **Retinal tear** | 130 (2.7) | 10 (1.5) | 0.070 |

**Legend:** IOFB, intraocular foreign body; VA, visual acuity; IQR, interquartile range; N, number.
