## Supplementary Table 7 for "Epidemiology, clinical features, and visual outcomes after intraocular foreign body removal: an IRIS® Registry (Intelligent Research in Sight) Analysis"

**Supplementary Table 7: Subgroup analysis of eyes with both pre-IOFB and post-IOFB VA.**

| **Predictor (Fixed effects)** | **Estimate (β)** | **95% CI** | **p-value** |
| --- | --- | --- | --- |
| (Intercept) | 0.91 | 0.77 – 1.04 | <0.001 |
| Time period (ref: Baseline) |  |  |  |
| 1-month post-IOFB | -0.21 | -0.26 – -0.16 | <0.001 |
| 2 months post-IOFB | -0.56 | -0.62 – -0.49 | <0.001 |
| 3 months post-IOFB | -0.57 | -0.64 – -0.50 | <0.001 |
| 4 months post-IOFB | -0.57 | -0.65 – -0.49 | <0.001 |
| 5 months post-IOFB | -0.59 | -0.68 – -0.51 | <0.001 |
| 6 months post-IOFB | -0.57 | -0.66 – -0.49 | <0.001 |
| 7 months post-IOFB | -0.58 | -0.67 – -0.49 | <0.001 |
| 8 months post-IOFB | -0.51 | -0.60 – -0.41 | <0.001 |
| 9 months post-IOFB | -0.55 | -0.65 – -0.45 | <0.001 |
| 10 months post-IOFB | -0.58 | -0.69 – -0.48 | <0.001 |
| 11 months post-IOFB | -0.61 | -0.72 – -0.49 | <0.001 |
| 12 months post-IOFB | -0.60 | -0.72 – -0.49 | <0.001 |
| 13 months post-IOFB | -0.64 | -0.76 – -0.52 | <0.001 |
| 14 months post-IOFB | -0.55 | -0.68 – -0.43 | <0.001 |
| 15 months post-IOFB | -0.59 | -0.72 – -0.45 | <0.001 |
| 16 months post-IOFB | -0.68 | -0.82 – -0.53 | <0.001 |
| 17 months post-IOFB | -0.72 | -0.87 – -0.57 | <0.001 |
| 18 months post-IOFB | -0.64 | -0.80 – -0.48 | <0.001 |
| Pre-IOFB VA |  |  |  |
| Pre-IOFB VA 0.5–1.0 (ref: VA < 0.5) | 0.53 | 0.37 – 0.68 | <0.001 |
| Pre-IOFB VA >1.0 (ref: VA < 0.5) | 0.92 | 0.78 – 1.07 | <0.001 |
| Sociodemographic factors |  |  |  |
| Age (reference: <65 years) | 0.01 | -0.03 – 0.04 | 0.876 |
| Sex: Female (ref: Male) | -0.01 | -0.12 – 0.10 | 0.944 |
| Race: Asian (ref: White) | 0.36 | -0.12 – 0.84 | 0.245 |
| Race: Black or African American (ref: White) | 0.33 | 0.10 – 0.56 | 0.014 |
| Race: Other (ref: White) | 0.14 | -0.42 – 0.70 | 0.803 |
| Race: Unknown (ref: White) | 0.03 | -0.22 – 0.27 | 0.932 |
| Ethnicity: Hispanic or Latino (ref: Not Hispanic or Latino) | 0.07 | -0.14 – 0.27 | 0.744 |
| Ethnicity: Unknown (ref: Not Hispanic or Latino) | -0.04 | -0.19 – 0.10 | 0.777 |
| Median income: USD 34,999 and below (ref: 35,000-74,999) | 0.00 | -0.44 – 0.43 | 0.985 |
| Median income: USD 75,000-149,999 (ref: 35,000-74,999) | 0.05 | -0.09 – 0.19 | 0.702 |
| Median income: USD 150,000 and above (ref: 35,000-74,999) | 0.14 | -0.26 – 0.53 | 0.729 |
| Median income: Unknown (ref: 35,000-74,999) | 0.13 | -0.43 – 0.69 | 0.803 |
| Clinical factors |  |  |  |
| IOFB location: Anterior and posterior segment (ref: anterior segment) | 0.06 | -0.54 – 0.67 | 0.932 |
| IOFB location: Posterior segment (ref: anterior segment) | -0.04 | -0.16 – 0.09 | 0.767 |
| Presenting complication: Endophthalmitis | 0.71 | 0.53 – 0.90 | <0.001 |
| Presenting complication: Hyphema | 0.36 | 0.15 – 0.58 | 0.003 |
| Presenting complication: Vitreous hemorrhage | 0.39 | 0.14 – 0.65 | 0.009 |
| Presenting complication: Retinal tear | 0.31 | -0.11 – 0.74 | 0.198 |
| Presenting complication: Retinal detachment | 0.51 | 0.30 – 0.72 | <0.001 |
| Presenting complication: Cataract | -0.07 | -0.31 – 0.16 | 0.753 |
| Interaction (Time period post-IOFB x pre-IOFB VA) |  |  |  |
| 1 month × VA 0.5–1.0 | 0.00 | -0.10 – 0.10 | 0.985 |
| 2 months × VA 0.5–1.0 | -0.01 | -0.13 – 0.11 | 0.944 |
| 3 months × VA 0.5–1.0 | -0.06 | -0.19 – 0.07 | 0.571 |
| 4 months × VA 0.5–1.0 | -0.01 | -0.15 – 0.14 | 0.975 |
| 5 months × VA 0.5–1.0 | -0.04 | -0.19 – 0.12 | 0.803 |
| 6 months × VA 0.5–1.0 | -0.16 | -0.32 – 0.00 | 0.096 |
| 7 months × VA 0.5–1.0 | -0.16 | -0.34 – 0.01 | 0.132 |
| 8 months × VA 0.5–1.0 | -0.18 | -0.35 – 0.00 | 0.096 |
| 9 months × VA 0.5–1.0 | -0.07 | -0.26 – 0.12 | 0.702 |
| 10 months × VA 0.5–1.0 | 0.00 | -0.19 – 0.19 | 0.985 |
| 11 months × VA 0.5–1.0 | -0.02 | -0.23 – 0.18 | 0.932 |
| 12 months × VA 0.5–1.0 | -0.12 | -0.33 – 0.09 | 0.448 |
| 13 months × VA 0.5–1.0 | -0.09 | -0.32 – 0.13 | 0.649 |
| 14 months × VA 0.5–1.0 | -0.04 | -0.27 – 0.19 | 0.866 |
| 15 months × VA 0.5–1.0 | 0.13 | -0.12 – 0.37 | 0.498 |
| 16 months × VA 0.5–1.0 | 0.06 | -0.20 – 0.33 | 0.803 |
| 17 months × VA 0.5–1.0 | 0.18 | -0.09 – 0.45 | 0.334 |
| 18 months × VA 0.5–1.0 | -0.10 | -0.39 – 0.19 | 0.729 |
| 1 month × VA >1.0 | 0.11 | 0.02 – 0.21 | 0.040 |
| 2 months × VA >1.0 | 0.27 | 0.15 – 0.38 | <0.001 |
| 3 months × VA >1.0 | 0.20 | 0.08 – 0.33 | 0.006 |
| 4 months × VA >1.0 | 0.15 | 0.01 – 0.29 | 0.064 |
| 5 months × VA >1.0 | 0.19 | 0.04 – 0.34 | 0.033 |
| 6 months × VA >1.0 | 0.19 | 0.04 – 0.34 | 0.033 |
| 7 months × VA >1.0 | 0.20 | 0.04 – 0.37 | 0.033 |
| 8 months × VA >1.0 | 0.18 | 0.01 – 0.35 | 0.086 |
| 9 months × VA >1.0 | 0.22 | 0.04 – 0.40 | 0.044 |
| 10 months × VA >1.0 | 0.22 | 0.03 – 0.41 | 0.046 |
| 11 months × VA >1.0 | 0.23 | 0.02 – 0.44 | 0.061 |
| 12 months × VA >1.0 | 0.17 | -0.05 – 0.38 | 0.237 |
| 13 months × VA >1.0 | 0.32 | 0.10 – 0.54 | 0.014 |
| 14 months × VA >1.0 | 0.17 | -0.07 – 0.40 | 0.287 |
| 15 months × VA >1.0 | 0.35 | 0.12 – 0.59 | 0.011 |
| 16 months × VA >1.0 | 0.34 | 0.09 – 0.60 | 0.021 |
| 17 months × VA >1.0 | 0.38 | 0.12 – 0.65 | 0.014 |
| 18 months × VA >1.0 | 0.38 | 0.09 – 0.68 | 0.026 |

**Legend**: VA, visual acuity; CI, confidence interval; IOFB, intraocular foreign body. Higher estimates (β) indicate worse visual acuity. Reference groups are baseline (time period 0), absent (for clinical factors), anterior (for IOFB location), White (for race), Not Hispanic or Latino (for ethnicity), and Male (for sex). Random Effects: Residual variance (σ²): 0.21; Intercept variance (eye ID) (τ₀₀): 0.41; Slope variance (time) (τ₁₁): 0.00; Correlation (intercept–slope) (ρ₀₁): 0.07; Intraclass correlation (ICC): 0.66. Model Fit: Number of eyes (clusters): 625; Observations: 8,438; Marginal R²: 0.375; Conditional R²: 0.787.
